# Japanese Psychiatrists’ Perceptions of Surgical Treatment for Treatment-Resistant Obsessive Compulsive Disorder: A Nationwide Survey

**DOI:** 10.64898/2025.12.11.25342024

**Authors:** Takashi Morishita, Shinsuke Kito, Kei Yamashiro, Daichi Konno, Hisatomi Arima, Hiroshi Abe, Masaki Iwasaki

## Abstract

**Aim:** Surgical interventions such as deep brain stimulation (DBS) have demonstrated efficacy for severe treatment-resistant obsessive–compulsive disorder (TR-OCD) internationally, but surgical treatment has yet to be implemented in Japan. One factor hindering progress is uncertainty regarding domestic clinical demand, so a nationwide survey of psychiatrists was conducted to clarify their perceptions of OCD surgery. We analysed the data with the aim of identifying factors associated with the acceptance of surgical interventions.

**Methods:** A web-based survey was conducted targeting board-certified psychiatrists in Japan. A total of 212 psychiatrists participated, responding to 10 multiple-choice questions assessing their awareness, perceived need, and expectations regarding DBS. Descriptive statistics, correlation analysis, and multiple linear regression models were used to examine associations between key variables.

**Results:** Perceived inadequacy of standard treatments alone for severe TR-OCD was strongly associated with perceived need for new treatment options including surgical interventions and with justifiability of surgical treatment. Clinical experience with OCD (Q3) showed a modest positive association with surgical acceptability, whereas greater experience with TR-OCD (Q4) corresponded to more cautious attitudes. Awareness of overseas surgical treatment (Q5) was associated with stronger recognition of treatment limitations and greater acceptance of surgical options.

**Conclusions:** Psychiatrists’ support for OCD surgery in Japan is primarily driven by perceived unmet clinical needs and awareness of international surgical practices. Enhancing education, addressing ethical concerns, and generating domestic clinical evidence may facilitate the responsible introduction of surgical treatment for TR-OCD.

## Introduction

Obsessive–compulsive disorder (OCD) is a chronic psychiatric disorder characterized by persistent obsessions and compulsions, leading to significant impairment of daily functioning and quality of life, with a prevalence of approximately 2.3%.^1, 2^ Standard treatments for OCD include selective serotonin reuptake inhibitors and cognitive behavioral therapy.^3^ However, a subset of individuals does not achieve sufficient symptom control with medical treatment and behavioral therapy and are considered to have treatment-resistant OCD (TR-OCD). For such cases, neurosurgical interventions including lesion therapy and deep brain stimulation (DBS) have been explored, particularly overseas.^4–6^ These treatments have been applied based on the theory of aberrant basal neurocircuitry involving the basal ganglia in OCD, and the theory has been widely accepted.^7–9^ Even though surgical treatment, especially DBS, being supported by clinical trials demonstrating its efficacy and safety,^6, 10, 11^ surgery for OCD has not yet been performed in Japan due to ethical issues and historical stigma.^12, 13^

Another important factor hindering the implementation of OCD surgery in Japan is the uncertainty regarding domestic demand. To obtain approval in Japan, clinical trials must demonstrate the efficacy of the surgical treatment, and there must be a certain level of demand to obtain funding for those studies. Therefore, understanding psychiatrists’ perspectives is crucial, but no large-scale survey has been conducted in Japan to assess their views on surgery as a potential treatment option for TR-OCD. To address this gap, we conducted a nationwide survey of board-certified psychiatrists in Japan to evaluate their awareness, perceived need, and expectations regarding OCD surgery. The findings of this survey shed light on the potential demand for OCD surgery among psychiatrists in Japan. The study aimed to identify key factors for gaining the understanding of psychiatrists, which is essential for developing strategies to advance the implementation of OCD surgery in Japan.

## Methods

### Study Design

This study was designed as a cross-sectional survey to assess psychiatrists’ perspectives on DBS as a treatment for TR-OCD in Japan. The survey was conducted using a web-based survey service provided by M3, Inc. (Tokyo, Japan), which enables rapid data collection from a large panel of medical professionals. The questionnaire was distributed online via m3.com, a major medical professional platform, and targeted board-certified psychiatrists. The initial target sample size was 200 respondents. The survey included multiple-choice and Likert-scale questions covering areas such as awareness of DBS, perceived need for surgical intervention, and expectations regarding its future implementation in clinical practice.

The participants were board-certified psychiatrists in Japan who met the eligibility criteria of holding certification from the Japanese Society of Psychiatry and Neurology and being actively engaged in clinical practice. The survey was designed to capture a wide range of clinical perspectives on DBS for TR-OCD. Recruitment via the m3.com platform ensured access to psychiatrists from various institutional backgrounds. A total of 212 psychiatrists completed the survey, with no additional exclusion criteria applied.

To assess the statistical precision of our sample, we estimated 95% confidence intervals for proportions observed in the survey. For a sample size of 200, the confidence interval for a response proportion of 50% is approximately ±7% (43%–57%), while for 10%, it is ±4% (6%–14%). This margin of error was considered adequate for the purposes of this study. No additional exclusion criteria were applied to ensure a broad spectrum of responses. The study protocol was approved by the Fukuoka University-Ethics Review Board (Approval Number: H25-563). As the survey was conducted with the aim of evaluating the clinical demand for surgical treatment, particularly DBS, the data were retrospectively analyzed after ethical approval was obtained, and the results were reported in accordance with the approved protocol. Written informed consent was waived by the ethics committee owing to the retrospective use of anonymized survey data and the absence of personally identifiable information.

### Survey Content

The survey consisted of 10 structured questions designed to assess psychiatrists’ awareness, perceived need, and expectations regarding surgical treatment for TR-OCD. The questionnaire was developed based on the existing literature and expert consultations to ensure its relevance to clinical practice. All questions were presented in a multiple-choice format, incorporating categorical responses and Likert-scale ratings. The full survey questionnaire, including all items and response options, is provided as Supplementary Material for reference.

The survey first collected demographic and professional background information, including psychiatrists’ board certification status and years since board certification (Q1), as well as the type of medical institution where they primarily work (Q2). The survey was designed such that respondents who reported not having board certification in Q1 were prevented from continuing to Q2 and beyond. The next questions examined participants’ clinical experience with OCD and TR-OCD, assessing the number of all OCD patients treated by each participant in the past year (Q3) and the number of patients with severe TR-OCD treated during the same period (Q4).

We evaluated participants’ awareness of overseas surgical treatments (e.g., DBS) for TR-OCD (Q5) and their perceived need for new treatment options, including neurosurgical interventions (Q6). Additionally, the survey assessed participants’ perceived inadequacy of standard treatments for TR-OCD (Q7). The final section of the survey focused on psychiatrists’ future outlook on the implementation of OCD surgery in Japan, exploring their expectations for regulatory approval and insurance coverage of surgical treatment (Q8), their opinion on the justifiability of surgical treatment options for severe TR-OCD (Q9), and their expectations regarding the incorporation of surgical treatments into future clinical guidelines (Q10).

### Statistical Analysis

Descriptive statistics were used to summarize participants’ responses, including frequency distributions, mean values, and standard deviations for each survey item. To assess relationships between variables, correlation analysis was conducted using Pearson’s correlation coefficient. Furthermore, multiple linear regression models were constructed to identify factors associated with the perceived need for new treatment options including surgery (Q6) and justifiability of surgical treatment options for severe TR-OCD (Q9). Independent variables included participants’ years since board certification (Q1), the number of all OCD patients treated by each participant (Q3), the number of patients with severe TR-OCD treated by each participant (Q4), and the perceived inadequacy of standard treatments alone for severe TR-OCD (Q7). To evaluate the influence of awareness of surgical treatment options overseas (Q5) on participants’ responses to the survey items, we conducted independent-samples t-tests (Welch’s t-test) comparing those who were aware of such options (Q5 = 1) with those who were not (Q5 = 2). The analysis was performed for all variables except Q2 (type of institution), which represents institutional characteristics and was excluded as a non-clinical factor.

A p-value of <0.05 was considered statistically significant. All statistical analyses were performed using EZR (Saitama Medical Center, Jichi Medical University, Saitama, Japan), which is a graphical user interface for R software (version 4.2.3; The R Foundation for Statistical Computing, Vienna, Austria).^14^

## Results

### Descriptive Statistics

The distribution of responses from a total of 212 board-certified psychiatrists for each survey item is summarized below. For years since board certification (Q1), 12.7% (n=27) had less than 5 years of experience, 16.5% (n=35) had 5–10 years, 22.6% (n=48) had 10–15 years, 29.7% (n=63) had 15–20 years, and 18.4% (n=39) had over 20 years. For the type of medical institution where the participants work (Q2), 8.0% (n=17) worked in university hospitals, 7.1% (n=15) in general hospitals with psychiatric wards, 5.2% (n=11) in general hospitals without psychiatric wards, 57.1% (n=121) in psychiatric hospitals, and 22.6% (n=48) in clinics or private practice.

For the number of all OCD patients treated by each participant in the past year (Q3), 44.3% (n=94) had treated fewer than 10 patients, 36.3% (n=77) treated 10–29, 12.7% (n=27) treated 30–49, 2.4% (n=5) treated 50–99, and 4.2% (n=9) treated 100 or more. The number of patients with severe TR-OCD treated by each participant (Q4) was 0 for 20.8% (n=44) of respondents, 1–4 for 49.5% (n=105), 5–9 for 18.9% (n=40), 10–19 for 6.1% (n=13), and 20 or more for 4.7% (n=10).

Regarding participants’ awareness of surgical treatments for TR-OCD (Q5), 50.9% (n=108) of respondents were aware of DBS as a surgical treatment for TR-OCD, while 49.1% (n=104) were not. When asked about the perceived need for new treatment options, including surgical interventions (Q6), 8.0% (n=17) considered them “very necessary,” 46.2% (n=98) “necessary,” 38.2% (n=81) were neutral, 6.6% (n=14) found them “unnecessary,” and 0.9% (n=2) considered them “completely unnecessary.”

In evaluating participants’ perceived inadequacy of standard treatments alone for severe TR-OCD (Q7), 9.4% (n=20) believed that TR-OCD patients frequently experience insufficient treatment effects, 48.1% (n=102) considered it “common,” 31.6% (n=67) were neutral, 9.4% (n=20) found it “rare,” and 1.4% (n=3) thought it was “very rare.” When asked about their expectations for regulatory approval and insurance coverage of surgical treatment (Q8), 4.7% (n=10) considered these “very likely,” 25.0% (n=53) considered these “likely,” 42.5% (n=90) were neutral, 21.7% (n=46) considered these “unlikely,” and 6.1% (n=13) considered these “very unlikely.”

Regarding the justifiability of surgical treatment options for severe TR-OCD (Q9), 9.9% (n=21) believed it should be “strongly considered,” 51.4% (n=109) thought it “should be considered,” 33.5% (n=71) were neutral, 4.7% (n=10) believed it “should not be considered,” and 0.5% (n=1) thought it “should never be considered.” Finally, concerning their expectations regarding the incorporation of surgical treatments into future clinical guidelines (Q10), 6.6% (n=14) considered it “very likely,” 31.6% (n=67) considered it “likely,” 45.3% (n=96) were neutral, 15.1% (n=32) considered it “unlikely,” and 1.4% (n=3) considered it “very unlikely.” These descriptive results are summarized in Figure 1.

**Figure 1.**
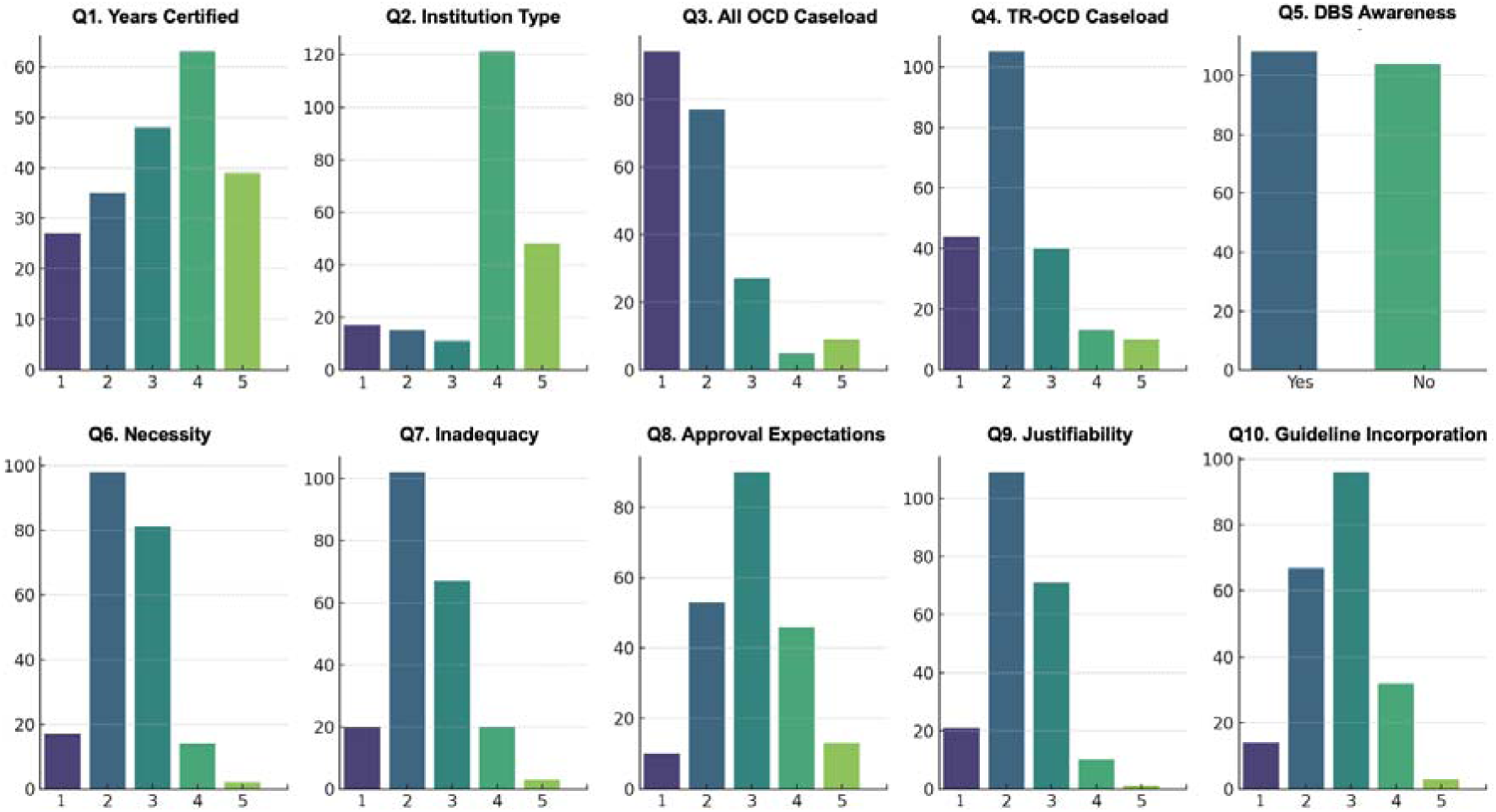
Distribution of responses to the questionnaires.

### Correlation Analysis

The Spearman correlation analysis demonstrated that psychiatrists who treated a greater number of patients with severe TR-OCD in the past year (Q4) tended to report a stronger perceived inadequacy of standard treatments alone for severe TR-OCD (Q7; r = – 0.38, p < 0.01) and a higher perceived need for new treatment options, including surgical interventions (Q6; r = –0.17, p = 0.02). The number of OCD patients treated in the past year (Q3) was strongly associated with the number of patients with severe TR-OCD treated (Q4; r = 0.62, p < 0.01), but showed only weak inverse relationships with attitudes toward surgical interventions (Q6, Q8–Q10). Importantly, perceived inadequacy of standard treatments alone for severe TR-OCD (Q7) was correlated with perceived need for new treatment options (Q6; r = 0.49, p < 0.01).

Furthermore, psychiatrists who more strongly perceived the need for new treatment options (Q6) or recognized the limitations of standard treatments (Q7) showed greater expectations for regulatory approval and insurance coverage of surgical treatment (Q8; r = 0.34 and 0.36, respectively; p < 0.01) and stronger support for the justifiability of surgical treatment options for severe TR-OCD (Q9; r = 0.40 and 0.44, respectively; p < 0.01). The expectation that surgical treatments will be incorporated into future clinical guidelines (Q10) was significantly associated with perceived need for new treatment options (Q6; r = 0.38, p < 0.01), Q7 (r = 0.44, p < 0.01), and particularly justifiability of surgical treatment (Q9; r = 0.57, p < 0.01), indicating that ethical acceptability is a key determinant of expectations regarding future implementation. These findings suggest that attitudes toward surgical interventions are driven by recognition of unmet clinical needs and direct experience with TR-OCD. These findings are summarized in Figure 2.

**Figure 2.**
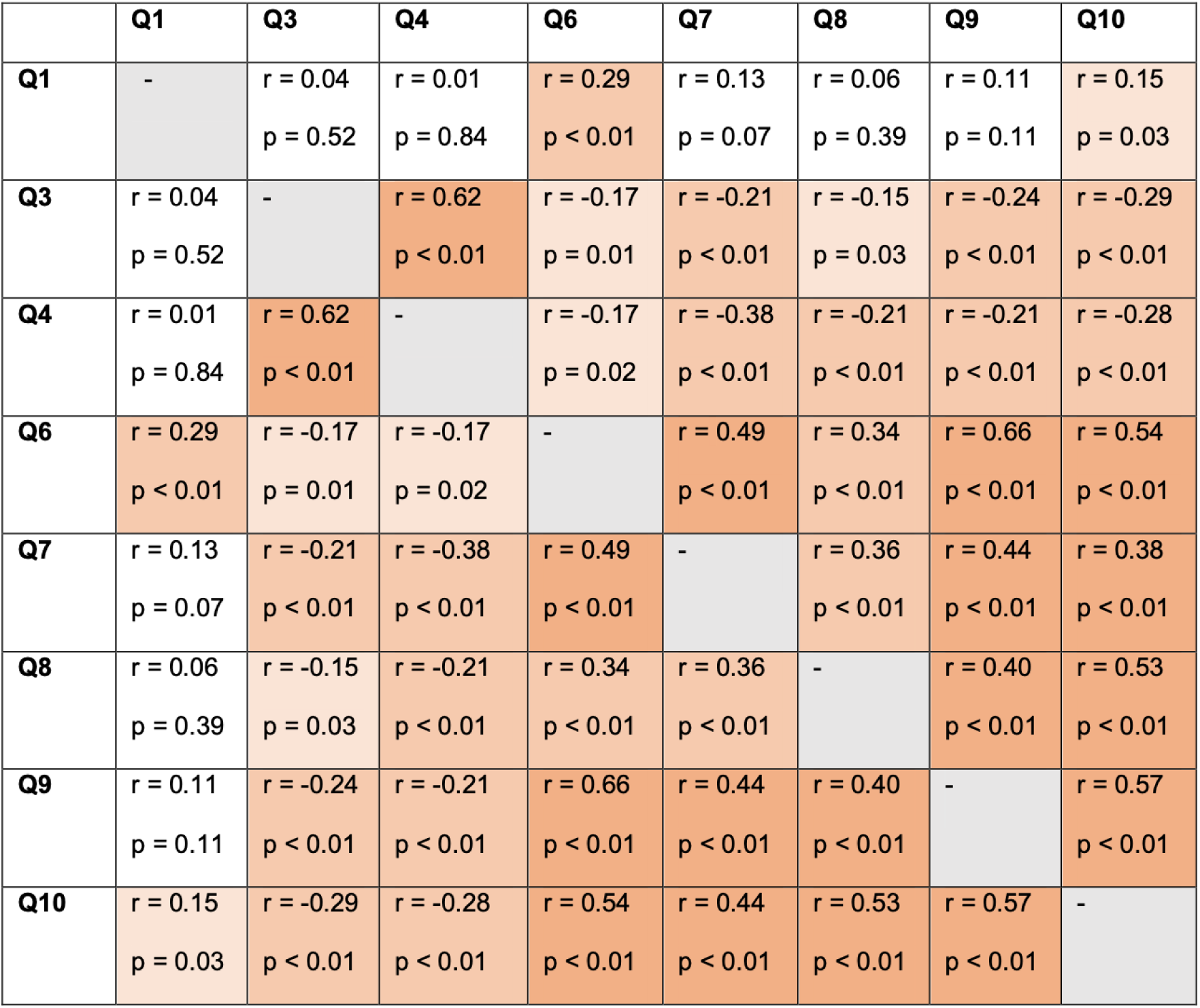
Summary correlation matrix among survey variables.

Cells are color-coded to visually represent the strength of each association: darker orange indicates stronger correlations, lighter orange indicates weaker positive correlations, and grey shading denotes non-significant relationships. Statistically significant correlations (p < 0.05) are highlighted.

### Regression Analysis

In the multiple linear regression analysis assessing factors associated with the perceived need for new treatment options, including surgical interventions (Q6), years since obtaining board certification (Q1; β = 0.14, p < 0.01) and perceived inadequacy of standard treatments alone for severe TR-OCD (Q7; β = 0.42, p < 0.01) were significant predictors. Additionally, although number of OCD patients treated in the past year (Q3; β = -0.10, p = 0.09) and number of patients with severe TR-OCD treated in the past year (Q4; β = 0.10, p = 0.12) showed non-significant associations. Since higher Q6 scores indicate a lower perceived need, these findings imply that more experienced clinicians tend to be more cautious regarding the introduction of surgical interventions and those recognizing greater limitations of standard are likely to new treatment options are necessary.

For the model evaluating the justifiability of surgical treatment options for severe TR-OCD (Q9), significant associations were observed for number of OCD patients treated in the past year (Q3; β = -0.17, p < 0.01) and perceived inadequacy of standard treatments alone for severe TR-OCD (Q7; β = 0.38, p < 0.01). In addition, although years since obtaining board certification (Q1; β = 0.06, p = 0.12) did not reach statistical significance, its positive direction suggests that clinicians with longer professional experience may be somewhat more reserved in justifying surgical treatment. Number of patients with severe TR-OCD treated in the past year (Q4; β = 0.08, p = 0.20) also showed a non-significant trend toward reduced justifiability. Overall, while greater exposure to OCD patients appears to enhance openness to surgical interventions, greater awareness of treatment limitations and professional seniority may contribute to more cautious or ethically conservative attitudes. These results are summarized in Table 1.

**Table 1.** Multiple linear regression analyses for factors associated with the perceived need for new treatment options (Q6) and the justifiability of surgical treatment options for severe TR-OCD (Q9).

| Dependent Variable | Independent Variable | $\beta$ | 95%CI | p-value | Interpretation |
| --- | --- | --- | --- | --- | --- |
| **Q6** | **Q1** | 0.14 | [0.07, 0.21] | < 0.01 | Longer clinical experience was linked to lower perceived need. |
|  | **Q3** | -0.10 | [-0.21, 0.02] | 0.09 | N.S. |
|  | **Q4** | 0.10 | [-0.03, 0.23] | 0.12 | N.S. |
|  | **Q7** | 0.42 | [0.31, 0.54] | < 0.01 | Psychiatrists who perceived standard treatments alone as insufficient were more likely to view new treatment options as necessary. |
| **Q9** | **Q1** | 0.06 | [-0.01, 0.13] | 0.12 | N.S. |
|  | **Q3** | -0.17 | [-0.28, -0.05] | < 0.01 | Psychiatrists treating more OCD patients were more likely to consider surgical options as appropriate. |
|  | **Q4** | 0.08 | [-0.04, 0.20] | 0.20 | N.S. |
|  | **Q7** | 0.38 | [0.26, 0.49] | < 0.01 | Psychiatrists who considered standard treatments insufficient were more likely to accept surgical treatment options. |

N.S., not significant.

### Independent-samples t-tests

Psychiatrists who were aware of overseas surgical treatment (e.g., DBS) for severe OCD (Q5) demonstrated a significantly greater perceived need for new treatment options, including surgical interventions (Q6, p = 0.01), higher perceived inadequacy of standard treatments alone for severe TR-OCD (Q7, p = 0.02), and stronger recognition of the justifiability of surgical treatment options for severe TR-OCD (Q9, p < 0.01). Awareness of surgical treatment was not associated with years since obtaining board certification (Q1), the number of OCD patients treated in the past year (Q3), or the number of patients with severe TR-OCD treated in the past year (Q4) (all p > 0.15). In addition, it did not affect expectations for regulatory approval and insurance coverage of surgical treatment (Q8) or expectations that surgical treatments will be incorporated into future clinical guidelines (Q10). These findings suggest that awareness of surgical treatment options overseas, rather than clinical experience or institutional background, plays a critical role in shaping clinicians’ attitudes toward the appropriateness of surgical intervention for TR-OCD. These findings are summarized in Table 2.

**Table 2.** Comparisons of question items according to awareness of overseas surgical treatment (Q5: Yes vs No).

| Variable | Mean<br>(Q5 = Yes) | Mean<br>(Q5 = No) | t-value | p-value | Interpretation |
| --- | --- | --- | --- | --- | --- |
| **Q1** | 3.19 | 3.30 | -0.59 | 0.56 | N.S. |
| **Q3** | 1.91 | 1.81 | 0.71 | 0.48 | N.S. |
| **Q4** | 2.34 | 2.14 | 1.44 | 0.15 | N.S. |
| **Q6** | 2.33 | 2.60 | -2.49 | 0.01 | Awareness associated with stronger perceived need. |
| **Q7** | 2.32 | 2.59 | -2.28 | 0.02 | Awareness associated with greater recognition of treatment limitations. |
| **Q8** | 2.99 | 3.00 | -0.07 | 0.94 | N.S. |
| **Q9** | 2.14 | 2.56 | -4.26 | < 0.01 | Awareness associated with stronger justification |
| **Q10** | 2.64 | 2.83 | -1.62 | 0.11 | N.S. |

N.S. = Not Significant.

## Discussion

The overall responses of psychiatrists in Japan were positive toward surgical treatment for TR-OCD despite the historical stigma. The present survey revealed that psychiatrists’ attitudes were primarily influenced by their recognition of the limitations of standard and, to a lesser extent, by their direct clinical experience with OCD patients. Greater perceived inadequacy of standard treatments alone for severe TR-OCD (Q7) was consistently associated with a higher perceived need for new treatment options (Q6) and greater acceptance of surgical interventions (Q9), as supported by both correlation and regression analyses. Although the number of patients with severe TR-OCD treated in the past year (Q4) showed a modest association with more supportive views, professional seniority (Q1) was inversely related to perceived need, suggesting a more cautious stance among experienced clinicians. Furthermore, awareness of surgical treatment options overseas (Q5) was associated with more favorable perspectives on clinical need and justifiability of surgical intervention. Overall, these findings highlight that acknowledgment of unmet treatment needs and awareness of existing surgical evidence are key drivers of support for the implementation of OCD surgery in Japan.

In Western countries, psychiatrists have shown cautious yet growing acceptance of DBS as a treatment for TR-OCD.^15^ Clinical studies suggest that direct experience with TR-OCD patients increases support for DBS, as psychiatrists who frequently encounter severe cases are more likely to consider neurosurgical interventions as a viable option.^16^ Additionally, long-term follow-up of patients treated with DBS for various psychiatric disorders, including OCD, has demonstrated sustained symptom improvement in a subset of patients, reinforcing its acceptance among clinicians.^10, 17^ Overseas, where DBS has been approved for select psychiatric indications, psychiatrists often view it as a viable treatment option.^17^ This may be attributed to well-established multidisciplinary screening processes, strict patient selection criteria, and regulatory frameworks that facilitate controlled clinical use.^18^

In general, the process of approving psychiatric indications for surgical treatments has also become more rigorous, with an emphasis on multidisciplinary ethical review boards and extensive patient screening.^10^ These developments suggest that while OCD surgery is a promising intervention, its adoption requires careful consideration of both clinical and ethical aspects as has frequently been addressed in the literature.^18–20^ Given that ethical and social concerns have been key barriers to psychiatric neurosurgery in Japan,^12^ fostering broader acceptance of OCD surgery requires not only clinical evidence but also structured discussions on long-term safety, regulatory oversight, and ethical frameworks to ensure responsible implementation.

To facilitate collaborative efforts by multidisciplinary teams including psychiatrists and neurosurgeons for the future implementation of surgical treatment options for TR-OCD in Japan, our study findings underscore the importance of targeted educational and policy strategies. First, increasing awareness of surgical treatment options (e.g., DBS) for severe OCD may help improve clinicians’ perceived need for new surgical treatments and acceptance of surgical interventions by psychiatrists. Second, deepening engagement with OCD specialists who recognize the inadequacy of standard treatment for severe TR-OCD may support the development of research initiatives and clinical protocols tailored to patients with unmet therapeutic needs.

Furthermore, economic factors play a crucial role in determining the feasibility of DBS integration into national healthcare systems. Unlike pharmacotherapy, surgical treatments such as DBS entails significant upfront costs for device implantation and long-term management, raising concerns about cost-effectiveness.^21^ Even though reimbursement policies vary widely across countries, with limited coverage for psychiatric indications,^22^ the favorable cost-effectiveness of OCD surgery has been reported by several groups.^23–25^ Similarly, the cost-effectiveness of DBS therapy for Parkinson’s disease patients in Japan has been reported.^13^ In addition to these encouraging reports, we believe that understanding psychiatrists’ perspectives, as examined in this study, serves as a valuable first step in informing future discussions on DBS policy and clinical guidelines.

While this study provides valuable insights into psychiatrists’ perceptions of DBS for TR-OCD in Japan, several limitations should be acknowledged. First, the study was conducted as a cross-sectional survey, which captures psychiatrists’ opinions at a single point in time. Attitudes toward OCD surgery may evolve as more clinical evidence emerges, and future longitudinal studies could better assess how perceptions change over time. Second, sampling bias may exist despite our efforts to recruit a diverse group of psychiatrists. The survey was conducted via m3.com, which, while widely used among medical professionals, may not fully represent all psychiatrists practicing in Japan. Additionally, psychiatrists with a particular interest in TR-OCD or advanced treatment options may have been more likely to respond, potentially influencing the results. Future studies should consider broader recruitment strategies, including professional society-based sampling, to further minimize potential bias. Third, the study relied on self-reported data, which may be subject to response bias. Psychiatrists’ stated views on DBS may not always reflect their actual clinical decision-making behaviors. Future research should also explore patient perspectives on DBS for TR-OCD, as public and patient acceptance plays a critical role in determining the feasibility of introducing new treatment modalities. Additionally, cost-effectiveness analyses and ethical discussions tailored to Japan’s healthcare system will be necessary to inform regulatory decisions and clinical guidelines.

## Conclusion

This nationwide survey demonstrated that Japanese psychiatrists generally recognize the limitations of conventional treatment strategies for severe TR-OCD and acknowledge the potential need for new treatment options, including surgical interventions. Support for OCD surgery was strongly influenced by perceived inadequacy of standard treatments alone for severe TR-OCD and awareness of overseas surgical approaches, whereas greater clinical experience and longer professional tenure were associated with more cautious attitudes. These findings suggest that improving awareness of international evidence and addressing ethical and social concerns may be critical to gaining broader clinical acceptance of surgical treatment. Further multidisciplinary discussion and prospective clinical studies are warranted to facilitate informed decision-making and advance implementation of surgical options for OCD in Japan.

## Data Availability

All data produced in the present study are available upon reasonable request to the authors.

## Acknowledgements

TM has received grant support from a Japan Society for the Promotion of Science (JSPS) Grant-in-Aid for Scientific Research (C) (grant number: 23K08555). KY has received grant support from a JSPS Grant-in-Aid for Early-Career Scientists (grant number: 25K19951) and from a Fukuoka University Research Grant (Grant number: GR2508). MI was supported by the Japan Agency for Medical Research and Development (AMED) (grant number: JP24wm0625407) and by the Intramural Research Grant of the National Center of Neurology and Psychiatry for Neurological and Psychiatric Disorders (grant number: 5-3).

## Author Contributions

TM contributed to the conceptualization, data gathering and analysis, and manuscript writing. SK, DK, and MI contributed to the conceptualization the study and manuscript writing. KY performed statistical analyses. HA (Hisatomi Arima) and HA (Hiroshi Abe) supervised the study and reviewed the manuscript.

