## Supplementary Material for "Japanese Psychiatrists’ Perceptions of Surgical Treatment for Treatment-Resistant Obsessive Compulsive Disorder: A Nationwide Survey"

Contents

Supplementary Text. English Version of the Survey Questionnaire.

**Supplementary Text. English Version of the Survey Questionnaire.**
This document contains the full text of the survey administered to Japanese psychiatrists, translated into English.

Q1
Do you hold a board certification from the Japanese Society of Psychiatry and Neurology? If so, how many years have passed since certification?
1. Less than 5 years
2. 5 to 10 years
3. 10 to 15 years
4. 15 to 20 years
5. More than 20 years
6. Not certified

Q2
What type of medical institution do you belong to?
1. University hospital (public/private)
2. General hospital with psychiatric ward
3. General hospital without psychiatric ward
4. Psychiatric hospital
5. Clinic or private practice
6. Other (please specify)

Q3
How many OCD patients have you treated in the past year?
1. Fewer than 10
2. 10 to 29
3. 30 to 49
4. 50 to 99
5. 100 or more

Q4
Among the OCD patients you treated in the past year, how many were treatment-resistant (i.e., did not sufficiently improve with standard pharmacotherapy and cognitive-behavioral therapy) and had severe symptoms?

Definition of severe: Causes significant impairment in daily functioning, requiring assistance from others, making commuting, studying, and daily life extremely difficult.
1. 0
2. 1 to 4
3. 5 to 9
4. 10 to 19
5. 20 or more

Q5
Are you aware that deep brain stimulation (DBS) and other surgical treatments are performed overseas for severe OCD?
1. Yes
2. No

Q6
To what extent do you feel that new treatment options, including surgical treatments, are necessary for patients with severe treatment-resistant OCD?
1. Strongly necessary
2. Necessary
3. Neutral
4. Not necessary
5. Strongly unnecessary

Q7
Concerning treatment-resistant OCD, what percentage of OCD patients do you believe do not respond adequately to standard treatments?
1. Very high
2. High
3. Neutral
4. Low
5. Very low

Q8
To what extent do you think surgical treatments for patients with severe treatment-resistant OCD will be approved and covered by insurance in Japan in the future?
1. Very likely

2. Likely
3. Neutral
4. Unlikely
5. Very unlikely

Q9
Considering ethical and social issues, to what extent do you believe that DBS and other surgical treatment options should be considered for treatment-resistant severe OCD patients?
1. Should be strongly considered
2. Should be considered
3. Neutral
4. Should not be considered
5. Should never be considered

Q10
To what extent do you think DBS and other surgical treatments will be incorporated into future OCD clinical guidelines and standard treatment systems?
1. Very likely
2. Likely
3. Neutral
4. Unlikely
5. Very unlikely
